## Appendix for "Covid-19 vaccine effectiveness against general SARS-CoV-2 infection from the omicron variant: A retrospective cohort study"

Lior Rennert, PhD

Department of Public Health Sciences, Clemson University

517 Edwards Hall

Clemson, SC, 29601

864-656-7434

### **Appendix 1. Statistical models for vaccine protection (Table 2)**

**Vaccine protection (across manufacturers):**

$h\left( t | V_{i},P_{i},\boldsymbol{X}_{i} \right)=h_{0}\left( t \right)\times\exp\left\{ \alpha\times V_{Pi}\left( t \right)+\beta\times V_{Fi}\left( t \right)+\gamma\times V_{Bi}(t)+\nu\times P_{i}+\boldsymbol{\eta}'\boldsymbol{X}_{i} \right\}$	(Model 1.1)

- Protection from full vaccination: $1-\exp\left\{ \beta\right\}$
- Protection from booster dose: $1-\exp\left\{ \gamma\right\}$
  - Increase in protection from booster dose: $\exp\left\{ \beta\right\}-\exp\left\{ \gamma\right\}$

where

- $V_{Pi}(t)$ = 1 if subject *i* is partially vaccinated at time *t*, and 0 otherwise (partially vaccinated = 14 days past first dose of mRNA-1273 or BNT162b2 and without second dose by time *t*)
- $V_{Fi}(t)$ = 1 if subject *i* is fully vaccinated at time *t*, and 0 otherwise (fully vaccinated = 14 days past second dose of mRNA-1273 or BNT162b2)
- $V_{Bi\left( t \right)}=1$ if subject *i* is boosted at time t, and 0 otherwise (booster = 7 days past booster dose of mRNA-1273 or BNT162b2)
- $P_{i}$=1 if subject *i* has previous SARS-CoV-2 infection occurring prior to the follow-up period, and 0 otherwise.
  - Note that $P_{i}$ remains time-invariant in this setting since it is not possible to have a previous infection occur during the follow-up period (since this is the event of interest and would therefore be classified as an infection during follow-up).

For employees, $\boldsymbol{X}_{i}$ = covariate vector for subject *i*, and includes age, race/ethnicity (categories defined in Table 1), gender (categories defined in Table 1), affiliation status (faculty/staff), self-reported presence of any of the following conditions: high blood pressure, heart disease, diabetes, overweight or obesity, kidney disease or dialysis, previous stroke or other neurological condition affecting my ability to cough, liver disease, or lung disease, self-reported use of tobacco or nicotine products, and number of SARS-CoV-2 tests since Fall 2020. For students, $\boldsymbol{X}_{i}$ also includes indicator for graduate student status subject, and affiliation status is (residential housing/non-residential housing).

**Vaccine protection (by manufacturer)**

$h\left( t | \boldsymbol{V}_{i},P_{i},\boldsymbol{X}_{i} \right)=h_{0}\left( t \right)\times\exp\left\{ \begin{aligned} \sum_{k=1:2} \alpha_{k}\times V_{P_{k},i}\left( t \right)+\sum_{l=1:2} \beta_{l}\times V_{F_{l},i}\left( t \right)+\sum_{j=1:2} \gamma_{j}\times V_{B_{j},i}\left( t \right) \\ +\nu\times P_{i}+\boldsymbol{\eta}'\boldsymbol{X}_{i} \end{aligned} \right\}$	(Model 1.2)

- Protection from full vaccination by mRNA-1273: $1-\exp\left\{ \beta_{1} \right\}$
- Protection from full vaccination by BNT162b2: $1-\exp\left\{ \beta_{2} \right\}$
  - Differences in vaccine effectiveness (2-dose series) between mRNA-1273 and BNT162b2 is evaluated by testing the null hypothesis $H_{0}: \beta_{1}=\beta_{2}$
- Protection from booster dose by mRNA-1273: $1-\exp\left\{ \gamma_{1} \right\}$
- Protection from booster dose by BNT162b2: $1-\exp\left\{ \gamma_{2} \right\}$
  - Differences in booster effectiveness between mRNA-1273 and BNT162b2 is evaluated by testing the null hypothesis $H_{0}: \gamma_{1}=\gamma_{2}$.

where

- $V_{P_{1},i}(t)$ = 1 if subject *i* is partially vaccinated from mRNA-1273 at time *t*, and 0 otherwise
- $V_{P_{2},i}(t)$ = 1 if subject *i* is partially vaccinated from BNT162b2 at time *t*, and 0 otherwise
- $V_{F_{1},i}(t)$ = 1 if subject *i* is fully vaccinated from mRNA-1273 at time *t*, and 0 otherwise
- $V_{F_{2},i}(t)$ = 1 if subject *i* is fully vaccinated from BNT162b2 at time *t*, and 0 otherwise
- $V_{B_{1},i}\left( t \right)$ = 1 if subject *i* is boosted from mRNA-1273 at time t, and 0 otherwise
- $V_{B_{2},i}\left( t \right)$ = 1 if subject *i* is boosted from BNT162b2 at time t, and 0 otherwise

**Vaccine protection for mRNA-1273/BNT162b2 sequence versus mix matched 2-dose/booster**

$h\left( t | \boldsymbol{V}_{i},P_{i},\boldsymbol{X}_{i} \right)=h_{0}\left( t \right)\times\exp\left\{ \begin{aligned} \sum_{k=1:2} \alpha_{k}\times V_{P_{k},i}\left( t \right)+\sum_{l=1:2} \beta_{l}\times V_{F_{l},i}\left( t \right)+\sum_{j=1:2} \gamma_{j}\times V_{B_{1j},i}\left( t \right) \\ \delta\times V_{B_{2},i}(t)+\nu\times P_{i}+\boldsymbol{\eta}'\boldsymbol{X}_{i} \end{aligned} \right\}$	(Model 1.3)

- Protection from complete mRNA-1273 sequence (i.e. 2-dose mRNA-1273 followed by mRNA-1273 booster): $1-exp\{\gamma_{1}\}$
- Protection from complete BNT162b2 sequence (i.e. 2-dose BNT162b2 followed by BNT162b2 booster): $1-exp\{\gamma_{2}\}$
- Protection from mix-matched booster: $1-exp\{\delta\}$
- Difference between mix-matched booster and complete mRNA-1273 sequence: $H_{0}:\delta=\gamma_{1}$
- Difference between mix-matched booster and complete BNT162b2 sequence: $H_{0}:\delta=\gamma_{2}$

where

- $V_{P_{1},i}(t)$ = 1 if subject *i* is partially vaccinated from mRNA-1273 at time *t*, and 0 otherwise
- $V_{P_{2},i}(t)$ = 1 if subject *i* is partially vaccinated from BNT162b2 at time *t*, and 0 otherwise
- $V_{F_{1},i}(t)$ = 1 if subject *i* is fully vaccinated from mRNA-1273 at time *t*, and 0 otherwise
- $V_{F_{2},i}(t)$ = 1 if subject *i* is fully vaccinated from BNT162b2 at time *t*, and 0 otherwise
- $V_{B_{11},i}\left( t \right)$ = 1 if subject *i* is boosted from complete mRNA-1273 sequence at time *t*, and 0 otherwise
- $V_{B_{12},i}\left( t \right)$ = 1 if subject *i* is boosted from complete BNT162b2 sequence at time *t*, and 0 otherwise
- $V_{B_{2},i}\left( t \right)$ = 1 if subject *i* has mix-matched booster at time *t*, and 0 otherwise

**Vaccine protection by previous SARS-CoV-2 infection history (across manufacturers)**

$h\left( t | V_{i},P_{i},\boldsymbol{X}_{i} \right)=h_{0}\left( t \right)\times\exp\left\{ \begin{aligned} \alpha\times V_{Pi}\left( t \right)+\beta\times V_{Fi}\left( t \right)+\gamma\times V_{Bi}(t)+\nu\times P_{i}+\delta_{1}\times V_{Pi}\left( t \right)\times P_{i} \\ +\delta_{2}\times V_{Fi}\left( t \right)\times P_{i}+\delta_{3}\times V_{Bi}\left( t \right)\times P\_i+\boldsymbol{\eta}'\boldsymbol{X}_{i} \end{aligned} \right\}$	(Model 1.4)

- Interaction between full vaccination and previous infection: $exp\{\delta_{2}\}$
- Interaction between boosted and previous infection: $exp\{\delta_{3}\}$

### **Appendix 2. Statistical models for waning vaccine protection**

**Vaccine protection (across manufacturers):**

$h\left( t | V_{i},P_{i},\boldsymbol{X}_{i} \right)=h_{0}\left( t \right)\times\exp\left\{ \begin{aligned} \alpha_{1}\times V_{Pi}\left( t \right)+\alpha_{2}\times V_{Pi}\left( t \right)\times T_{{V_{P}}_{i}}+\beta_{1}\times V_{Fi}\left( t \right)+\beta_{2}\times V_{Fi}\left( t \right)\times T_{{V_{F}}_{i}}+ \\ \gamma_{1}\times V_{Bi}\left( t \right)+\gamma_{2}\times V_{Bi}\left( t \right)\times T_{V_{Bi}}+\nu_{1}\times P_{i}+\nu_{2}\times P_{i}\times T_{P_{i}}+\boldsymbol{\eta}'\boldsymbol{X}_{i} \end{aligned} \right\}$	(Model 2.1)

- Full vaccination: hazard ratio for change in monthly risk: $\exp\left\{ \beta_{2}\times30 \right\}$
- Booster dose: hazard ratio for change in monthly risk: $\exp\left\{ \gamma_{2}\times30 \right\}$

where

- $T_{V_{P_{i}}}$ is days between date of partial vaccination and time *t*
- $T_{V_{F_{i}}}$ is days between date of full vaccination and time *t*
- $T_{V_{B_{i}}}$ is days between date of booster and time *t*
- $T_{P_{i}}$ is days between date of previous SARS-CoV-2 infection and time *t*

**Vaccine protection (by manufacturer)**

$h\left( t | \boldsymbol{V}_{i},P_{i},\boldsymbol{X}_{i} \right)=h_{0}\left( t \right)\times\exp\left\{ \begin{aligned} \sum_{k=1:2} \alpha_{1k}\times V_{P_{k},i}\left( t \right)+\sum_{k=1:2} \alpha_{2k}\times V_{P_{k},i}\left( t \right)\times T_{V_{P_{i},k}} \\ +\sum_{l=1:2} \beta_{1l}\times V_{F_{l},i}\left( t \right)+\sum_{l=1:2} \beta_{2l}\times V_{F_{l},i}\left( t \right)\times T_{V_{C_{i},l}}+\sum_{j=1:2} \gamma_{1j}\times V_{B_{j},i}\left( t \right)+ \\ \sum_{j=1:2} \gamma_{2j}\times V_{B_{j},i}\left( t \right)\times T_{V_{B_{i},j}}+\nu_{1}\times P_{i}+\nu_{2}\times P_{i}\times T_{P_{i}}+\boldsymbol{\eta}'\boldsymbol{X}_{i} \end{aligned} \right\}$	(Model 2.2)

- Differences in change in monthly risk between mRNA-1273 and BNT162b2 among fully vaccinated: $H_{0}: \beta_{21}=\beta_{22}$
- Differences in change in monthly risk between mRNA-1273 and BNT162b2 among fully vaccinated: $H_{0}: \gamma_{21}=\gamma_{22}$

where

- $T_{V_{P_{1},k}}$ is days between date of first dose of mRNA-1273 and time *t*
- $T_{V_{P_{2},k}}$ is days between date of first dose of BNT162b2 and time *t*
- $T_{V_{F_{1},k}}$ is days between date of second dose of mRNA-1273 and time *t*
- $T_{V_{F_{2},k}}$ is days between date of second dose of BNT162b2 and time *t*
- $T_{V_{B_{1},k}}$ is days between date of booster dose of mRNA-1273 and time *t*
- $T_{V_{B_{2},k}}$ is days between date of booster dose of BNT162b2 and time *t*

**Vaccine protection (across manufacturers) with common decline over time**

$h\left( t | V_{i},P_{i},\boldsymbol{X}_{i} \right)=h_{0}\left( t \right)\times\exp\left\{ \alpha\times V_{Bi}\left( t \right)+\beta\times T_{Vi}+\nu_{1}\times P_{i}+\nu_{2}\times P_{i}\times T_{Pi}+\boldsymbol{\eta}'\boldsymbol{X}_{i} \right\}$	(Model 2.3)

- Time-adjusted risk of SARS-CoV-2 infection: $exp\{\alpha\}$

where

- $T_{Vi}=\left\{ \begin{aligned} days between date of full vaccination and time t, if subject i fully vaccinated \\ days between date of boosted and time t, if subject i \mathrm{boosted} \end{aligned} \right.$

This model is restricted to individuals who were fully vaccinated or boosted at the start of follow-up.

- Due to restricting the sample to fully vaccinated individuals who are not booster eligible during the follow-up period (i.e., received their second dose less than 5.25 months prior to end of follow-up), $V_{Bi}\left( t \right)$ is fixed for each subject *i* in this model (i.e., $V_{Bi}\left( t \right)=V_{Bi})$

### **Appendix 3. Additional Tables and Figures**

**Table S1**. STROBE checklist for cohort studies.

|  | Item No | Recommendation | Main Text Page |
| --- | --- | --- | --- |
| **Title and abstract** | 1 | (*a*) Indicate the study’s design with a commonly used term in the title or the abstract | Abstract |
|  |  | (*b*) Provide in the abstract an informative and balanced summary of what was done and what was found | Abstract |
| Introduction | | |  |
| Background/rationale | 2 | Explain the scientific background and rationale for the investigation being reported | Introduction |
| Objectives | 3 | State specific objectives, including any prespecified hypotheses | Introduction |
| Methods | | |  |
| Study design | 4 | Present key elements of study design early in the paper | Methods (‘Study Design and Population’) |
| Setting | 5 | Describe the setting, locations, and relevant dates, including periods of recruitment, exposure, follow-up, and data collection | Methods (‘Study Design and Population’, ‘SARS-CoV-2 Testing’, ‘Vaccination Status’), Figure 1 |
| Participants | 6 | (*a*) Give the eligibility criteria, and the sources and methods of selection of participants. Describe methods of follow-up | Methods (‘Study Design and Population’, ‘SARS-CoV-2 Testing’, ‘Vaccination Status’) |
|  |  | (*b*) For matched studies, give matching criteria and number of exposed and unexposed | Methods (‘Propensity Score Matching’) |
| Variables | 7 | Clearly define all outcomes, exposures, predictors, potential confounders, and effect modifiers. Give diagnostic criteria, if applicable | Methods (‘Statistical Analyses’), Table 1A/B |
| Data sources/ measurement | 8* | For each variable of interest, give sources of data and details of methods of assessment (measurement). Describe comparability of assessment methods if there is more than one group | Methods (‘Study Design and Population’, ‘SARS-CoV-2 Testing’, ‘Vaccination Status’) |
| Bias | 9 | Describe any efforts to address potential sources of bias | Methods (‘Study Design and Population’, ‘Vaccination Status’, ‘Propensity Score Matching’, ‘Statistical Analyses’) |
| Study size | 10 | Explain how the study size was arrived at | Figure 1 |
| Quantitative variables | 11 | Explain how quantitative variables were handled in the analyses. If applicable, describe which groupings were chosen and why | Methods (‘Propensity Score Matching’, ‘Statistical Analyses’), Table 1A/B |
| Statistical methods | 12 | (*a*) Describe all statistical methods, including those used to control for confounding | Methods (‘Propensity Score Matching’, ‘Statistical Analyses’), Appendix 1-2 |
|  |  | (*b*) Describe any methods used to examine subgroups and interactions | Methods (‘Statistical Analyses’), Appendix 1-2 |
|  |  | (*c*) Explain how missing data were addressed | NA |
|  |  | (*d*) If applicable, explain how loss to follow-up was addressed | Methods (‘Statistical Analyses’). Individuals who did not test positive for SARS-CoV-2 during the follow-up period were censored at their last negative test date. |
|  |  | (*e*) Describe any sensitivity analyses | NA |
| Results | | |  |
| Participants | 13* | (a) Report numbers of individuals at each stage of study—eg numbers potentially eligible, examined for eligibility, confirmed eligible, included in the study, completing follow-up, and analysed | Results (paragraphs 1-3), Figure 1 and Table 1A/B |
|  |  | (b) Give reasons for non-participation at each stage |  |
|  |  | (c) Consider use of a flow diagram |  |
| Descriptive data | 14* | (a) Give characteristics of study participants (eg demographic, clinical, social) and information on exposures and potential confounders | Results (paragraphs 1-2), Table 1A/B |
|  |  | (b) Indicate number of participants with missing data for each variable of interest | NA |
|  |  | (c) Summarise follow-up time (eg, average and total amount) | Results (‘Vaccine Effectiveness’, paragraphs 1-2) |
| Outcome data | 15* | Report numbers of outcome events or summary measures over time | Results (paragraphs 1-2), Table 1A/B, Table 2 |
| Main results | 16 | (*a*) Give unadjusted estimates and, if applicable, confounder-adjusted estimates and their precision (eg, 95% confidence interval). Make clear which confounders were adjusted for and why they were included | Results (‘Vaccination Effectiveness’, paragraphs 1-3), Table 2, Figure 2 |
|  |  | (*b*) Report category boundaries when continuous variables were categorized | NA |
|  |  | (*c*) If relevant, consider translating estimates of relative risk into absolute risk for a meaningful time period | NA |
| Other analyses | 17 | Report other analyses done—eg analyses of subgroups and interactions, and sensitivity analyses | Results (‘Vaccination Effectiveness’, paragraph 4) |
| Discussion | | |  |
| Key results | 18 | Summarise key results with reference to study objectives | Discussion (paragraphs 1-2) |
| Limitations | 19 | Discuss limitations of the study, taking into account sources of potential bias or imprecision. Discuss both direction and magnitude of any potential bias | Discussion (paragraphs 5-6) |
| Interpretation | 20 | Give a cautious overall interpretation of results considering objectives, limitations, multiplicity of analyses, results from similar studies, and other relevant evidence | Discussion (paragraph 7) |
| Generalisability | 21 | Discuss the generalisability (external validity) of the study results | Discussion (paragraphs 3-6) |
| Other information | | |  |
| Funding | 22 | Give the source of funding and the role of the funders for the present study and, if applicable, for the original study on which the present article is based | Acknowledgements |

*Give information separately for exposed and unexposed groups.

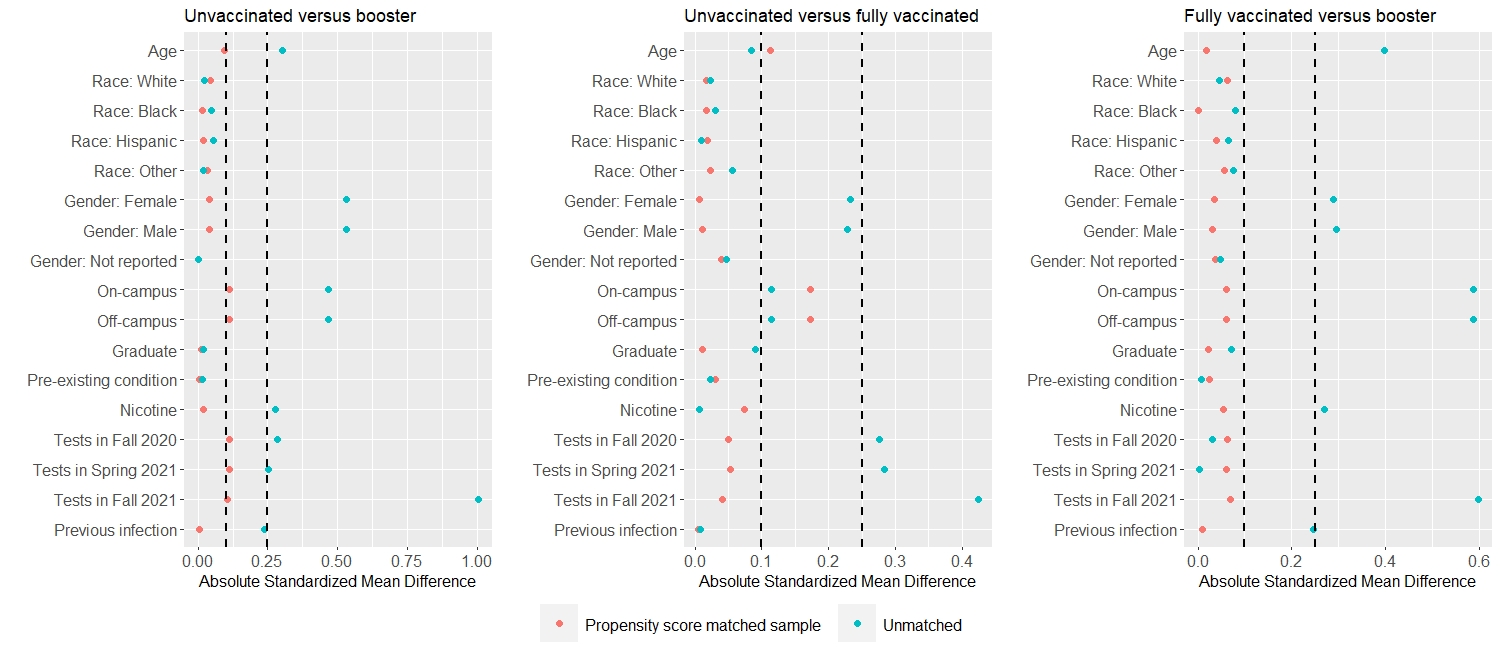

**Figure S1A.** Absolute standardized mean difference between unvaccinated versus booster (left) and unvaccinated versus fully vaccinated (right) for baseline covariates before and after propensity score matching among students. Vertical lines represent the thresholds of 0.1 and 0.25.

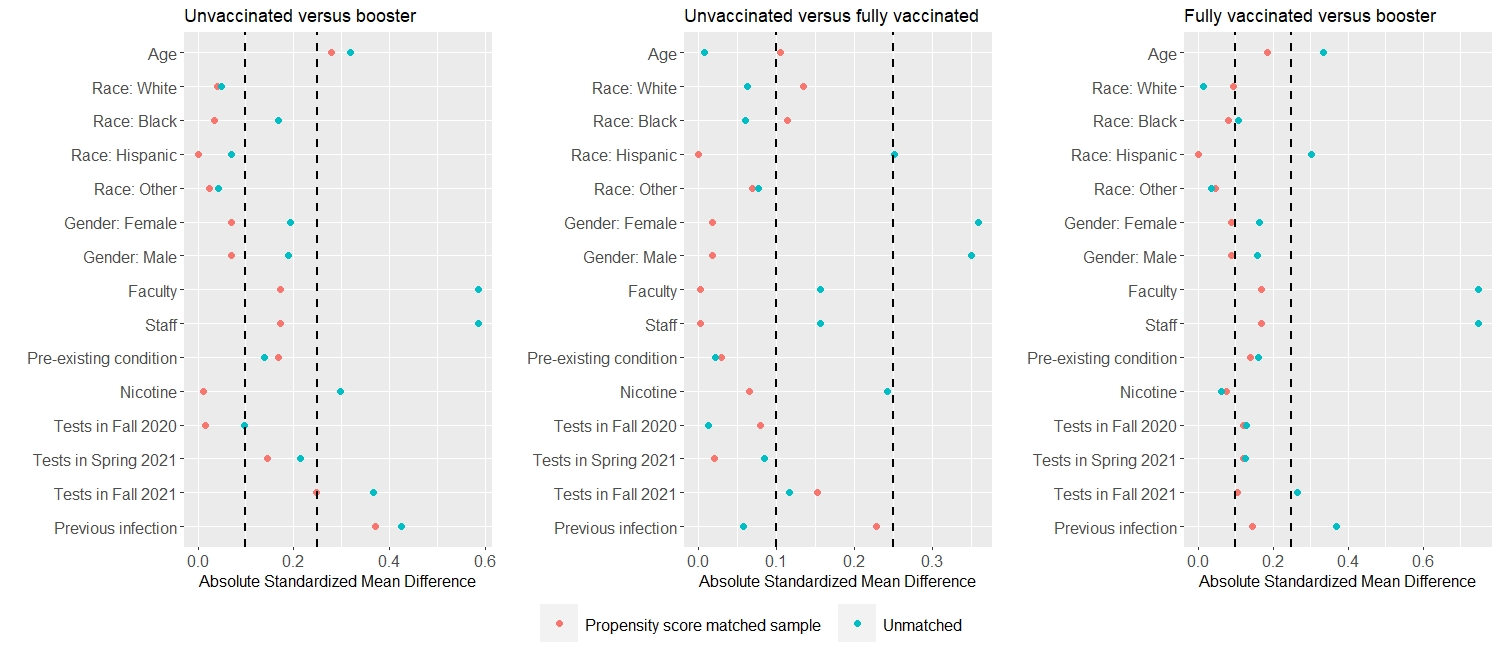

**Figure S1B.** Absolute standardized mean difference between unvaccinated versus booster (left) and unvaccinated versus fully vaccinated (right) for baseline covariates before and after propensity score matching among employees. Vertical lines represent the thresholds of 0.1 and 0.25.

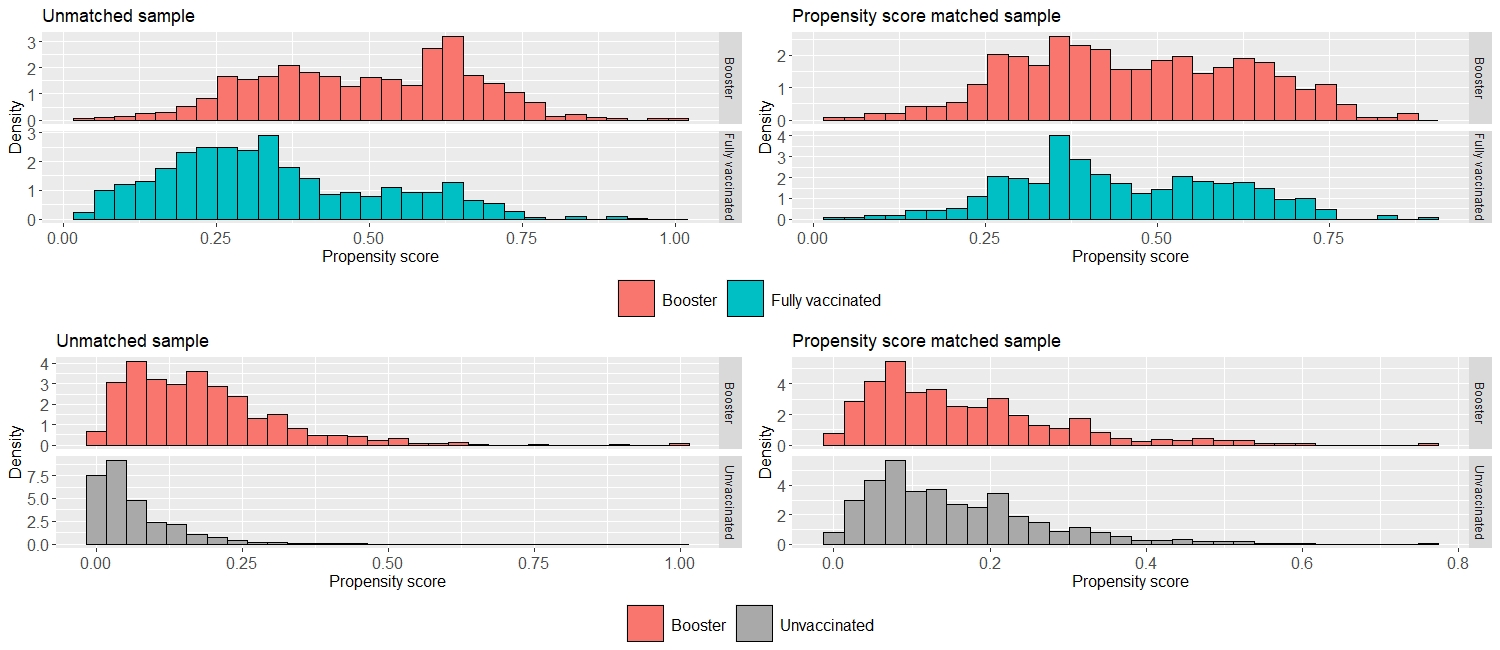

**Figure S2A.** Histogram of propensity scores of the unmatched sample and the matched sample for students. The common reference group (boosted) is represented in red. Unvaccinated and fully vaccinated are represented in grey and green, respectively.

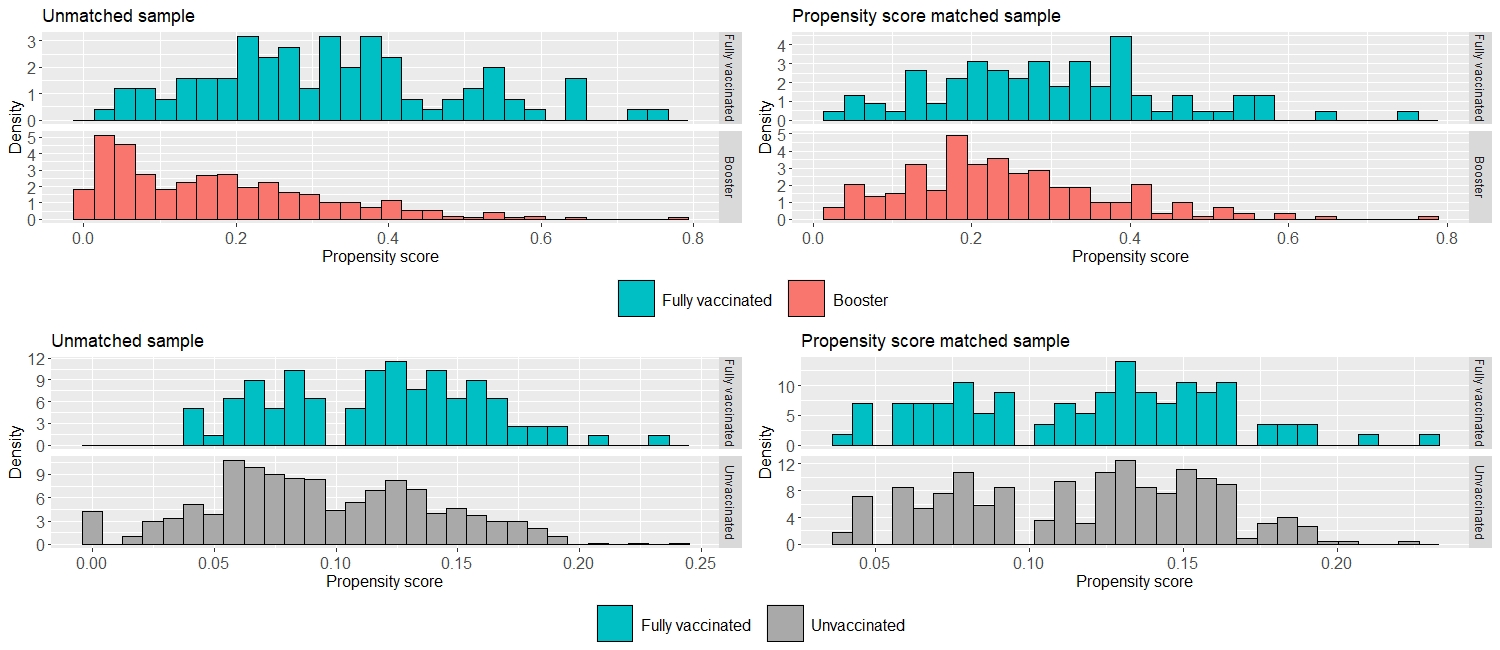

**Figure S2B.** Histogram of propensity scores of the unmatched sample and the matched sample for employees. The common reference group (fully vaccinated) is represented in green. Unvaccinated and booster are represented in grey and red, respectively.

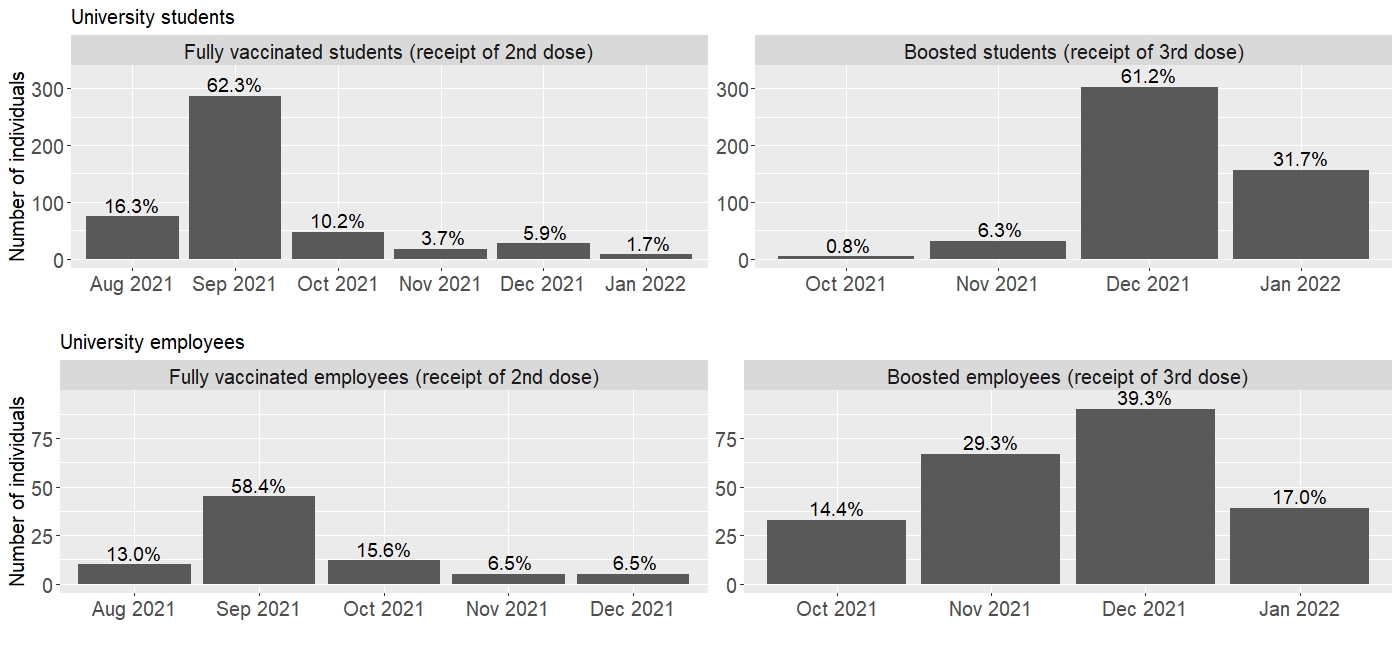

**Figure S3.** Distribution of receipt of the 2nd dose and the booster dose by calendar month for university students and employees in the matched sample.
